# Dementia diagnosis and prevalence in the Lothian Birth Cohort 1936 using medical data linkage

**DOI:** 10.1101/2022.11.18.22282515

**Authors:** Donncha S. Mullin, Lucy E. Stirland, Emily Buchanan, Catherine-Anne Convery, Simon R. Cox, Ian J. Deary, Cinzia Giuntoli, Holly Greer, Danielle Page, Elizabeth Robertson, Susan D. Shenkin, Anna Szalek, Adele Taylor, Georgina Weatherdon, Tim Wilkinson, Tom C. Russ

## Abstract

**Background:** The Lothian Birth Cohort 1936 (LBC1936) is a longitudinal study of ageing with well-characterised assessments, but until now, it has relied on self-report or proxies for dementia outcomes. This report describes a framework for clinical dementia ascertainment and its implementation. We report the prevalence of all-cause dementia and dementia subtypes in 865 individuals aged 70 years and older from the LBC1936.

**Methods:** Electronic Health Records (EHR) of all participants were reviewed, and relevant information was extracted to form case vignettes for everyone with any record of cognitive dysfunction. The EHR data sources include hospital and clinic letters, general practitioner and hospital referrals, prescribed medications, imaging and laboratory results. Death certificate data were obtained separately. Clinician assessments were performed when there was concern about a participant’s cognition. A diagnosis of probable dementia, possible dementia, or no dementia was agreed upon by a consensus diagnostic review board, comprised of a multidisciplinary team of clinical dementia experts who reviewed case vignettes and clinician assessment letters. For those with probable dementia, a subtype was also determined, where possible. Finally, we compared the agreement between our newly ascertained dementia outcomes with the existing self-reported dementia outcomes.

**Results:** The EHR review identified 163 out of 865 (18.8%) individuals as having cognitive dysfunction. At the consensus diagnostic review board, 118/163 were diagnosed with probable all-cause dementia, a prevalence of 13.6%. Age-specific dementia prevalence increased with age from 0.8% (65-74.9 years) to 9.93% (85-89.9 years). Prevalence rates for women were higher in nearly all age groups. The most common subtype was dementia due to Alzheimer disease (49.2%), followed by mixed Alzheimer and cerebrovascular disease (17.0%), dementia of unknown or unspecified cause (16.1%), and dementia due to vascular disease (8.5%). Self-reported dementia outcomes were positive in only 17.8% of ascertained dementia outcomes.

**Conclusions:** Dementia outcomes have been clinically ascertained in the LBC1936 using a robust systematic approach that closely aligns with diagnosing dementia in practice. This provides useful detailed outcomes for further analyses of LBC1936 to allow exploration of lifecourse predictors of dementia.

## Introduction

Dementia is a major and growing global public health crisis.[1] Dementia research is crucial for informing present and future demand for dementia care services.[2] As the number of people with dementia increases globally, obtaining accurate dementia prevalence rates based on valid and robust dementia ascertainment is crucial to guide health system planning and to inform research decisions. Epidemiological studies require robust dementia outcomes, ideally in well-characterised longitudinal cohorts, to allow the identification of lifecourse predictors of dementia, and to produce meaningful results to inform policy and clinical practice.

This report describes the derivation of a robust, clinically-derived dementia outcome in an important longitudinal cohort study, the Lothian Birth Cohort 1936 (LBC1936). We also describe prevalence rates of all-cause dementia and subtypes in this cohort. The LBC1936 study was designed as a study of healthy ageing; no members of the LBC1936 had a diagnosis of dementia when the study began (mean age 70 years).

## Methods

### Participants

This study used data from the LBC1936 (https://www.ed.ac.uk/lothian-birth-cohorts), described in detail elsewhere.[3–5] In summary, participants living in the Lothian region of Scotland (which includes Edinburgh), most of whom had completed an intelligence test aged 11 years, were recruited in 2004, at mean age 69.5 years (*n*=1091). At initial recruitment, none reported a diagnosis of dementia. They have been followed up every three years since, at mean ages 72.5 years (*n*=866), 76.3 years (*n*=697), 79.3 years (*n*=550) and 82 years (*n*=431). The sixth wave of data collection is currently underway, and a seventh is planned. All participants are white, and the sex split is approximately equal. At each wave, participants undergo a core battery of cognitive testing, including measures of reasoning, processing speed, executive function, and memory. In addition, a detailed medical history, blood tests, physical measures, structural magnetic resonance imaging (MRI; age 72.5 onwards) are collected at each wave. The neuropsychological battery performed as part of LBC1936 testing includes the Mini-Mental State Exam (MMSE), logical memory 1 & 2, verbal fluency, National Adult Reading Test, Weschler Test of Adult Reading, Test of Premorbid Functioning, digit symbol coding, backward digit span, simple and four-choice reaction time, block design, verbal paired associates, spatial span, symbol search, matrix reasoning, verbal paired associates delay, and inspection time.[4] Symptoms of depression and anxiety are measured using the Hospital Anxiety and Depression Screen.

We excluded participants who did not consent to data linkage to their medical records. LBC1936 participants were asked for their consent to access medical records from wave 2 onwards, so participants who attended wave 1 and did not return for a subsequent wave were excluded from our study.

### Dementia ascertainment process

Our diagnostic procedure followed a previously validated process[6] with the additional step of a clinical assessment at home, where indicated, on a selection of our cohort. As illustrated in Figure 1, there were three phases: (i) Electronic Health Record (EHR) review plus death certificate data, (ii) home visit clinician assessments, and (iii) consensus review board meeting.

**Figure 1:**
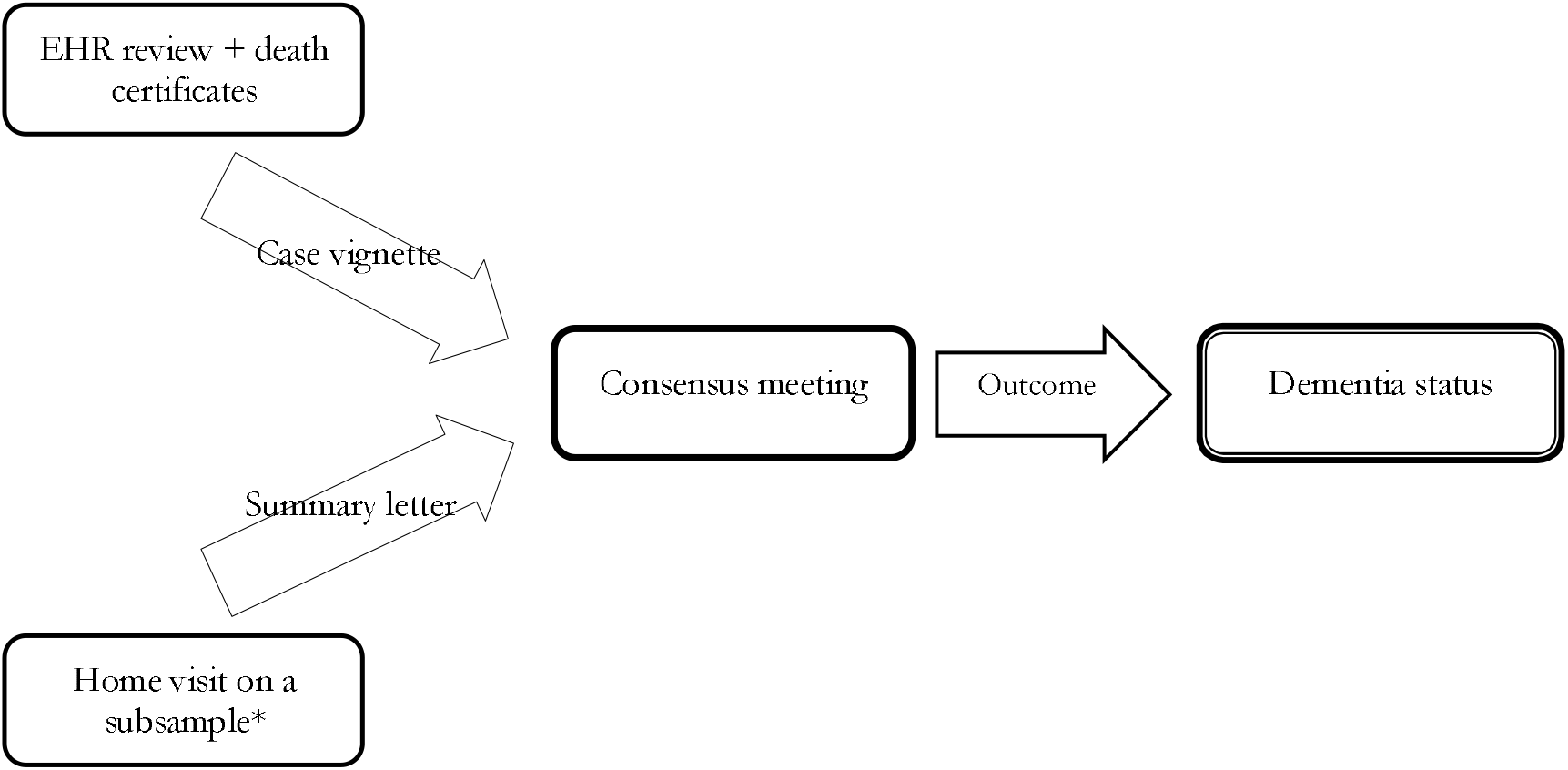
Overview of the dementia diagnostic process Note: EHR, Electronic Health Record *only a small subsample of participants had information available at the consensus review meeting following a home visit assessment. Doctor home visits were requested for several reasons, explained in the section ‘Home visits’.

#### Phase 1: Electronic Health Record (EHR) review

A team of psychiatrists specialising in Old Age Psychiatry (“EHR team”) reviewed the EHR of every consenting LBC1936 participant. All information was accessed and stored within the secure National Health Service computer system. An EHR protocol was produced by the group to ensure a standardised and systematic approach for each participant (Additional File 1).

The EHR for each participant was located using the patient’s Community Health Index (CHI) number, a unique health identifier used in NHS Scotland. Since 2014, all healthcare records within NHS Lothian (the health board covering Edinburgh and surrounding areas) including psychiatric records have been stored on the EHR as full-text letters, records of referrals from primary to secondary/tertiary care services, hospital discharge letters including medications, and results of laboratory and radiological investigations. Before 2014, general and psychiatric records were held on separate systems, but all records were subsequently incorporated into the TrakCare system. Death certificate data is available for all deceased LBC1936 participants via record linkage. This was checked for each participant at the diagnostic review board (see Phase 3, below).

##### Case vignettes

The psychiatrist who reviewed the EHR created anonymised extracts of relevant information for the diagnostic review board meeting for any participant with evidence of cognitive dysfunction or a diagnosis of dementia. This work was completed on 17^th^ April 2022.

Participants with upcoming NHS services investigations or assessments, such as brain imaging or memory clinic assessments, were flagged in the case vignettes to make the diagnostic review board aware. The EHRs of these flagged case vignettes were checked for updated information at the diagnostic review board.

#### Phase 2: Home visits

Doctor home visits were requested for several reasons: when cognitive impairment or decline was noted by LBC research staff during routine LBC1936 wave 6 testing (in comparison to test scores in prior waves); when a new diagnosis of dementia was self-reported to the LBC research team; or when the LBC researcher had concerns that the participant might have dementia. Wave 6 testing was ongoing at the time of our study. Before participating in Wave 6 of the study, LBC1936 participants were informed that they would be invited to have a home visit if there was a substantial decline in their cognitive scores or if they had already been diagnosed with dementia; participants provided written consent when attending their Wave 6 cognitive testing appointment.

During the home visit, an experienced Old Age psychiatrist performed a detailed clinical assessment. This included a thorough interview with the participant and informant, where available, to gather a complete medical history. Cognitive testing was completed using the Addenbrooke’s Cognitive Examination-III[7] and a physical examination, allowing the completion of the Modified Hachinski Ischaemic Scale.[8] The clinician also reviewed the participant’s medical records including investigations (laboratory results, brain imaging), clinic letters, and prescribed medications. They then wrote to the participant’s general practitioner detailing the outcome of the assessment and, if necessary, referred them for further assessment within the NHS. These letters were available for review by the consensus diagnostic review board and were considered alongside the case vignettes.

#### Phase 3: Consensus Diagnostic Review Board

The consensus group consisted of experienced dementia experts from Old Age Psychiatry (AS, CG, DM, LS, TR), Geriatric Medicine (SS), and Neurology (TW). We discussed each participant flagged as having either possible or probable dementia. The final date for this phase was 18^th^ August 2022. We agreed on whether the available evidence supported a diagnosis of dementia and determined the subtype of dementia, where possible. Depending on the strength of the evidence, both the diagnosis and subtype were deemed either ‘probable’ or ‘possible’. The criteria used for probable and possible diagnoses are shown in Table 1 (derived from a validated process[6]). Any disagreement was resolved through discussion. Any individual identified as having dementia but where there was insufficient evidence to make a subtype diagnosis was classified as an ‘unclear’ subtype. Differential diagnoses were made according to the ICD-11 criteria.[9]

**Table 1:** Criteria for probable and possible diagnoses utilised by the consensus team

| Probable Dementia | Possible Dementia |
| --- | --- |
| ANY of the following (without opposing evidence from same/other source): | ANY of the following (without opposing evidence from same/other source): |
| - dementia diagnosis on death certificate (any part)<br>- dementia diagnosed on clinical review (ICD-11/DSM-5)<br>- dementia diagnosis in electronic health records<br>- ICD-11 criteria for dementia diagnosis met by data within any existing records | - recorded cognitive impairment on death certificate<br>- cognitive impairment/decline recorded in notes, but incomplete evidence to meet ICD-11 diagnostic criteria<br>- possibility of dementia recorded in notes but no formal diagnosis/ incomplete evidence to meet ICD-11 diagnostic criteria |

Note: ICD-11, International Classification of Disease - Eleventh Edition; DSM-5, Diagnostic and Statistical Manual of Mental Disorders - Fifth Edition; LBC1936, Lothian Birth Cohort 1936.

The earliest date of any diagnosis was recorded. When only the month was available, we selected the middle of the month. Where only the year was available, 2^nd^ July was selected as the estimated middle of the year.

Participants flagged as having impending memory assessments or investigations, as described in Phase 1, were revisited at the consensus meeting, and any new information was also considered.

To minimise the risk of misclassification, any inconsistencies between data sources were considered on a case-by-case basis. If there was reliable and consistent evidence for dementia in one source (e.g., Psychiatry clinic letter), but not another (e.g., death certificate), it was assumed the participant had dementia. Where there was contradictory evidence of similar reliability from two sources, further evidence was sought from other sources, and a consensus was reached. If it was impossible to obtain further evidence, participants with contradictory evidence were classified as possible rather than probable dementia.

We arranged NHS clinical follow-ups for participants newly diagnosed with dementia in our study. We calculated the approximate number of person-hours each ascertainment phase took, to guide researchers considering replicating our methods in other cohorts. Approximately 469 person-hours were required to ascertain dementia in this cohort (Additional File 2), the majority required for phase 1 (400 hours).

### Analysis

First, we calculated the prevalence of all-cause probable dementia, i.e., the proportion of the study sample that, at some point in their life or between the ages of approximately 70 and 86 years (if they are still alive), developed dementia. We compared the basic characteristics of the participants with and without probable dementia. Second, we calculated the age-stratified dementia prevalence by removing those with dementia from the numerator when they died and removing all those who died from the denominator. Third, we calculated the age-stratified dementia incidence rate. We report five-year age groupings, but we pooled the two groups 65 to 69.9 years and 70 to 74.9 years to preserve anonymity due to sample distribution. Finally, we calculated the proportions of the probable dementia subtypes.

We compared the self-reported dementia outcomes at each study wave to the ascertained dementia outcomes (Figure 4). We recorded which sources contributed information for each probable dementia diagnosis (Figure 2). The information sources were categorised as EHR, clinical assessments at home, death certificates, and brain imaging. We noted if dementia was recorded on any part of the death certificate. For those participants who underwent more than one brain imaging modality, we noted the most detailed modality (e.g., MRI if the participant had had both CT and MRI brain scans). We considered brain imaging results from both NHS clinical settings and LBC1936 scans. Statistical analysis was performed using R version 4.0.2.[10] Code is openly available on GitHub.[11]

**Figure 2:**
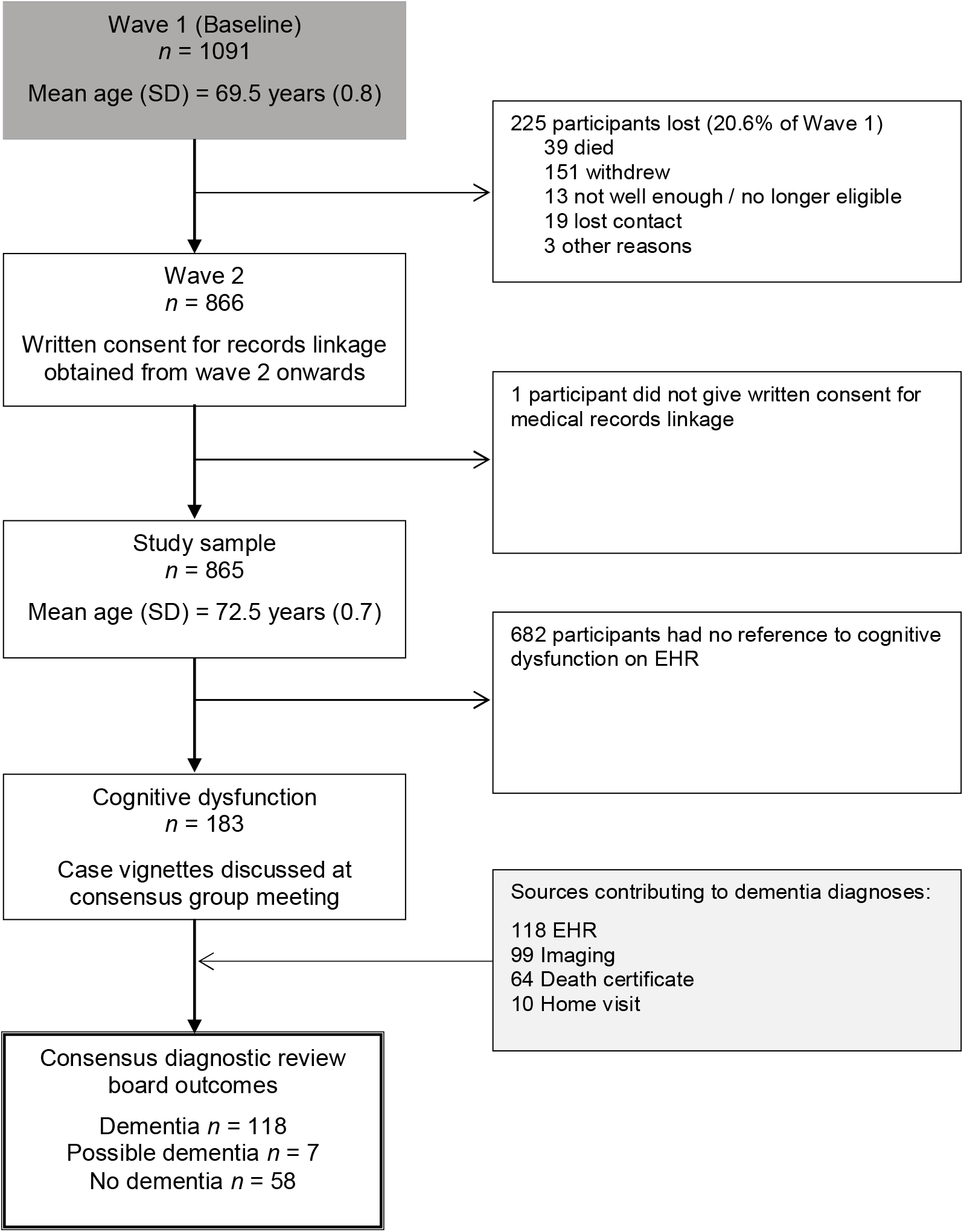
Participant flowchart, data sources contributing to dementia diagnoses, and consensus diagnostic review board outcomes. Note: SD, Standard Deviation; EHR, Electronic Health Record.

## Results

### Participants

Wave 2 of the LBC1936 had 866 participants. One participant did not consent to their data being linked to their medical records. Accordingly, we included 865 participants in our analysis. Of these, 163 participants (18.8%) were flagged as having cognitive dysfunction following the EHR review and/or home visit. The anonymised case vignettes derived from the EHR, along with home visit clinical assessment letters for 10 participants, formed the basis of the discussion at the consensus group meeting. We illustrate the flow of participants, the data sources contributing to dementia diagnoses, and the consensus diagnostic review board outcomes in Figure 2.

### Dementia prevalence

In this cohort of older adults who were free from dementia at study inception, we found that 13.6% (118/865) of participants met the criteria for a diagnosis of probable all-cause dementia between the ages of approximately 70 and 86 years old. Their basic demographics and IQ scores at age 11 are presented in Table 2.

**Table 2:** Demographics of the participants with and without probable dementia

|  |  | No Dementia<br>(n=747) | Dementia<br>(n=118) |
| --- | --- | --- | --- |
| Sex | Female (%) | 362 (48.5) | 55 (46.6) |
|  | Male (%) | 385 (51.5) | 63 (53.4) |
| Education, years | Mean (SD) | 10.8 (1.1) | 10.6 (1.1) |
| Age 11 IQ | Mean (SD) | 101.0 (15.0) | 98.5 (17.2) |
| Previous occupation | Manual | 148 (20.2) | 27 (23.5) |
|  | Non-manual | 586 (79.8) | 88 (76.5) |
| Marital Status | Married | 523 (70.0) | 92 (78.0) |
|  | Not married | 224 (30.0) | 26 (22.0) |

Note: IQ, Intelligence Quotient.

In addition to the 118 people with probable dementia, seven participants were diagnosed with possible dementia. Of these seven, six were deceased making further testing impossible.

### Age-stratified dementia prevalence

The prevalence of dementia for the age group 65 to 74.9 years was 0.8% rising to 9.47% for the age group 80-84.9 years. There was only a slight increase to 9.93% in the 85-89.9 years age group as the mean age of dementia diagnosis in this group was only 85.49 years. We pooled the two groups 65 to 69.9 years and 70 to 74.9 years to preserve anonymity due to sample distribution. The prevalence rates for women were higher in nearly all age groups. At the time of the consensus meeting, 321 of 865 participants had died; 64 of these had dementia. Thus, 54 participants with dementia were alive at the time of our study. They had a mean (SD) age of 86 (0.8) years, and the sex divide was approximately even. As individuals with dementia died, they were removed from our prevalence calculation for later age groups. Table 3 presents the age-stratified dementia prevalence in our study, both pooled and grouped by sex.

**Table 3:** Age-stratified dementia prevalence.

| Age group,<br>years | Alive with dementia |  |  |  |  |  |  |  |
| --- | --- | --- | --- | --- | --- | --- | --- | --- |
|  | Alive<br>*N*(%) | Prevalence<br>(per 1000) | Total<br>(*N*) | Men<br>(*N*) | Women<br>(*N*) | Total<br>(%) | Men<br>(%) | Women<br>(%) |
| 65-74.9 | 839 (96.9%) | 8.3 | 7 | 4 | 3 | 0.8% | 0.9% | 0.7% |
| 75-79.9 | 736 (85.1%) | 44.8 | 33 | 15 | 18 | 4.5% | 4.1% | 4.9% |
| 80-84.9 | 570 (65.9%) | 94.7 | 54 | 25 | 29 | 9.5% | 9.3% | 9.7% |
| 85-89.9^ | 544 (62.9%) | 99.3 | 54 | 22 | 32 | 9.9% | 8.7% | 11.0% |

Note: ^At the time of our study, the mean age of participants with dementia in the 85-89.9 years group is only 85.5 years old. This explains the relatively low number of new diagnoses in this group.

### Age-stratified dementia incidence

Table 4 presents the incident dementia diagnoses over five years, distributed across age and sex categories.

**Table 4:** Age-stratified dementia incidence

| Age group,<br>years | New dementia diagnoses |  |  |  |  |  |  |
| --- | --- | --- | --- | --- | --- | --- | --- |
|  | Incidence<br>Rate (per<br>1000) | Total<br>(*N*) | Men (*N*) | Women<br>(*N*) | Total (%)* | Men (%) | Women<br>(%) |
| 65-74.9 | 8.3 | 7 | 4 | 3 | 0.8% | 0.9% | 0.7% |
| 75-79.9 | 48.9 | 36 | 20 | 16 | 4.9% | 5.4% | 4.4% |
| 80-84.9 | 107.0 | 61 | 32 | 29 | 10.7% | 11.9% | 9.7% |
| 85-89.9^ | 23.9 | 13 | 6 | 7 | 2.4% | 2.4% | 2.4% |
| **Total** | - | **118** | **63** | **55** | - | - | - |

Note: \*Of the live participants in the age group.
^At the time of our study, the mean age of participants with dementia in the 85-89.9 years group is only 85.5 years old. This explains the relatively low number of new diagnoses in this group.

### Dementia subtypes

The distribution of the 118 probable dementia outcomes by subtype was as follows: dementia due to Alzheimer disease (49.2%), mixed Alzheimer and cerebrovascular disease (17.0%), vascular disease (8.5%), Lewy body disease (3.4%), dementia due to psychoactive substances (1.7%), diseases classified elsewhere (e.g., Creutzfeldt-Jakob Disease, Parkinson’s; 4.2%), and dementia of unknown or unspecified cause (16.1%). Table 5 presents the subtype diagnoses in detail. Figure 3 illustrates the main subtype groupings.

**Table 5:**
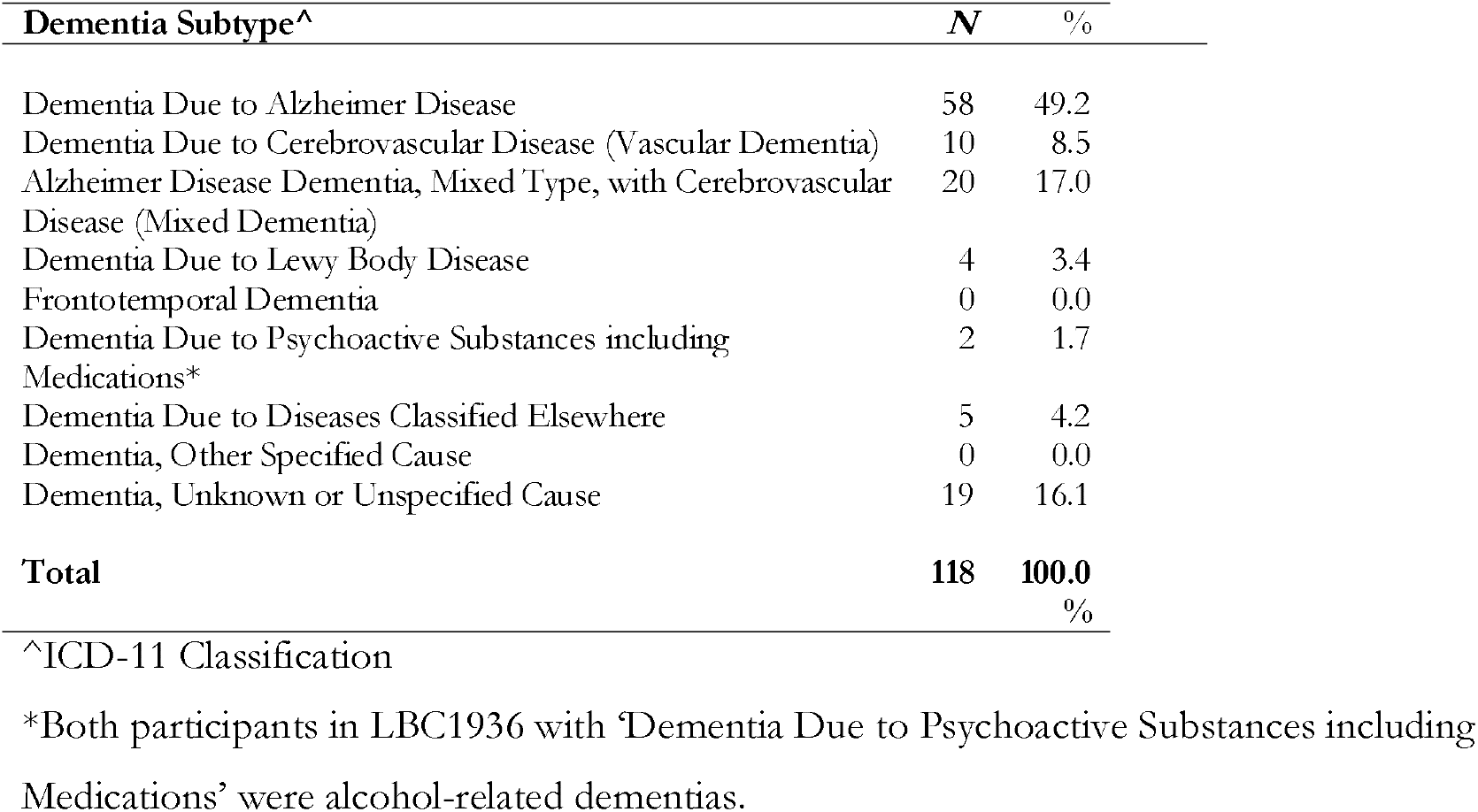
Distribution of dementia outcomes by subtype, in detail

**Figure 3:**
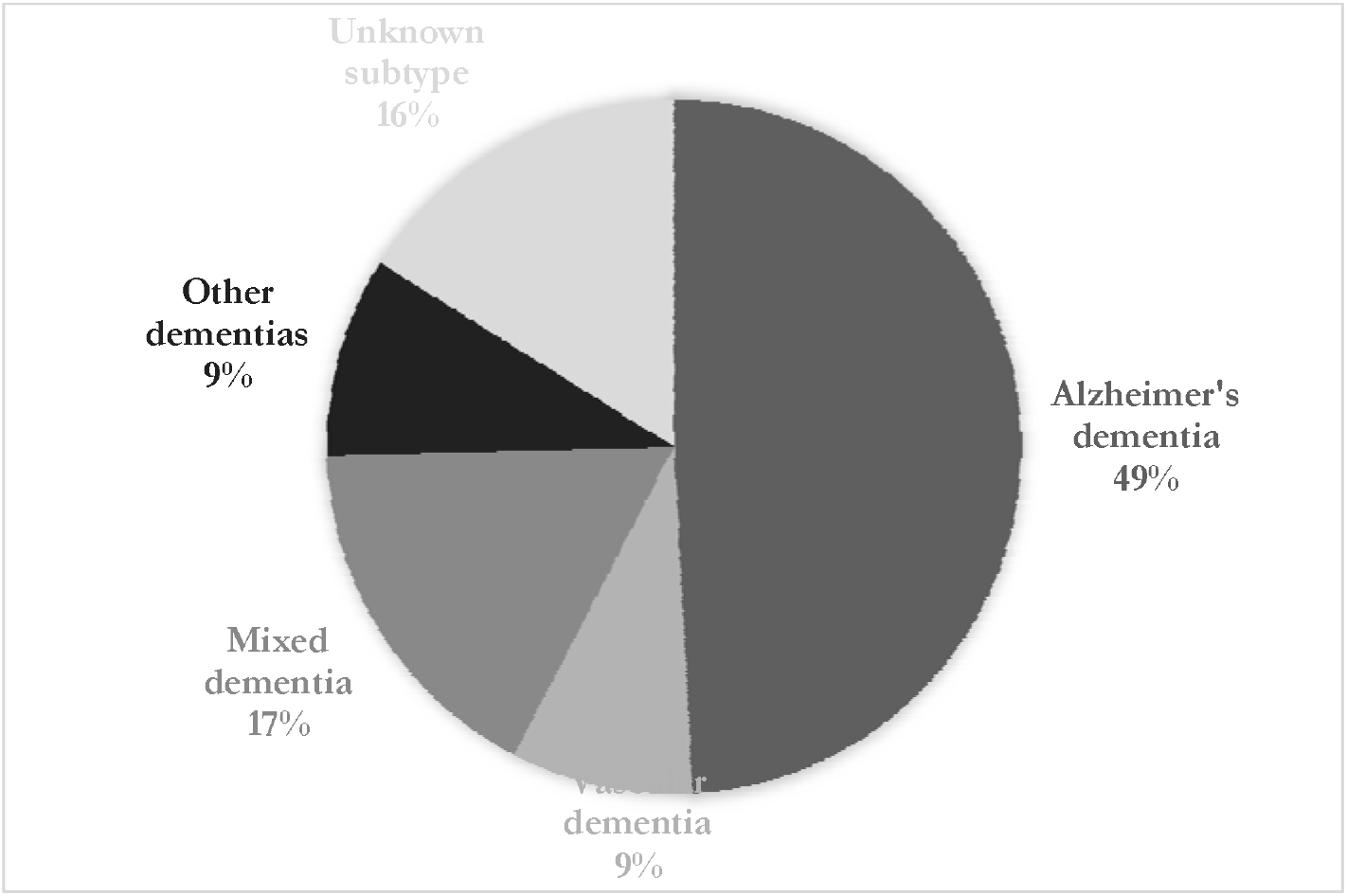
Distribution of main dementia subtype groups Note: “Other dementias” contains those due to Lewy Body Disease, psychoactive substances, and diseases classified elsewhere (precise proportions are presented in Table 5).

### Comparing self-reported and ascertained dementia outcomes

Figure 4 illustrates the large difference between the number of self-reported and ascertained dementia outcomes. Of the 118 ascertained dementia outcomes, only 21 had ever self-reported dementia. One participant who self-reported dementia did not have dementia ascertained. The self-reported dementia outcome in LBC1936, therefore, has a sensitivity of 17.8% and a specificity of 98.9%, when assigning the ascertained dementia outcomes as the gold standard.

**Figure 4:**
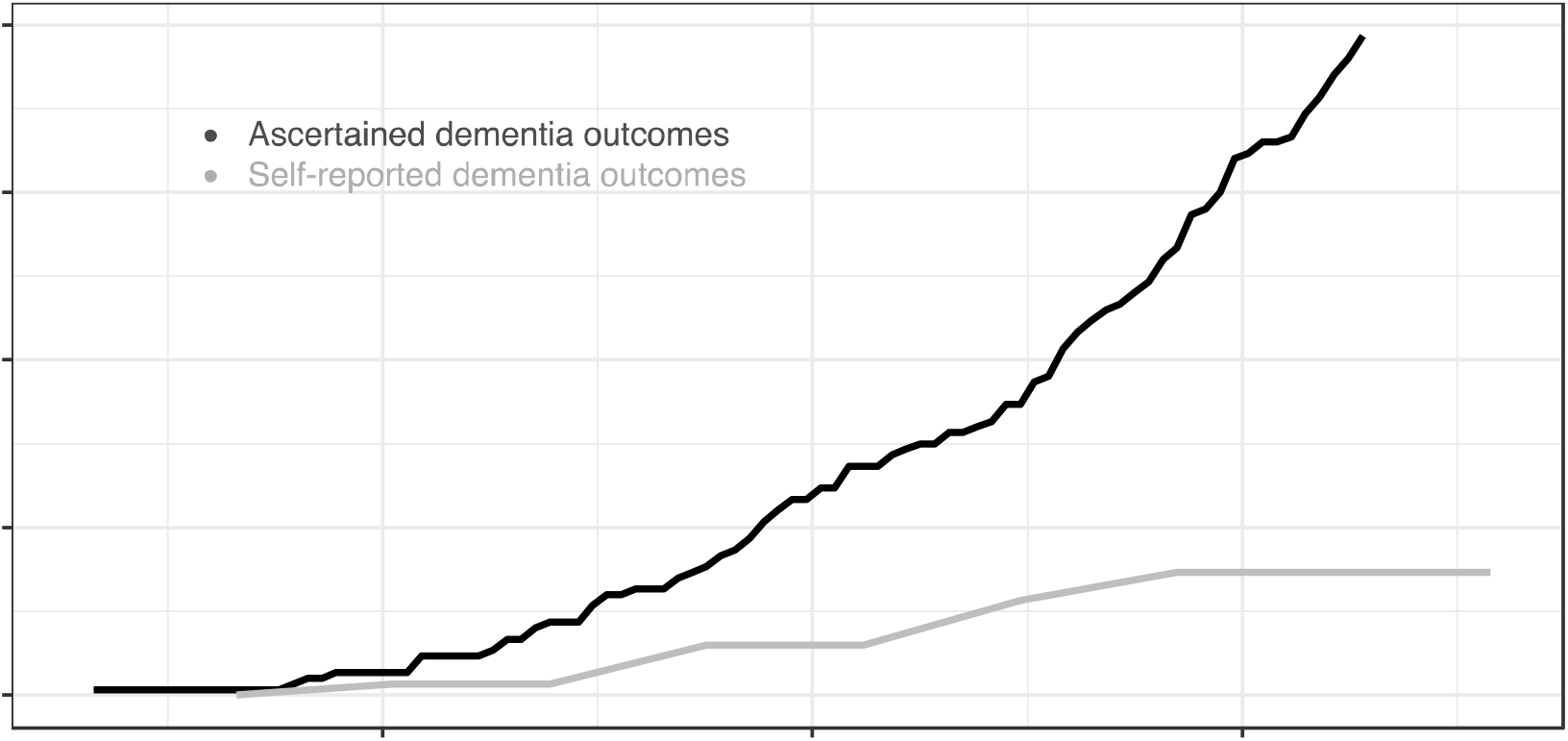
Comparison of self-reported and ascertained dementia outcomes Note: due to attrition, the number tested (i.e., asked about their dementia status) reduces at each wave, whereas access to electronic health records is not affected by attrition.

### Data sources contributing to probable dementia diagnoses

Of the 118 probable dementia diagnoses, 43 had an MRI brain scan (37%), 53 had a CT brain scan (45%), and three (2.5%) had another scan such as single-photon emission computerised tomography (SPECT) scan, Dopamine Transporter (DaT) Scan, or Positron emission tomography (PET). Nineteen (16%) of those with a dementia diagnosis did not have any brain imaging. As stated in the methods, we recorded only the most detailed scan a participant received, as this was given precedence during the consensus group meeting. Of the 64 participants with dementia who died, a diagnosis of dementia was recorded on the death certificate of 47 (73.4%). Information from home visits contributed to 10/118 dementia diagnoses. This information is illustrated in Figure 2.

## Discussion

We have ascertained dementia outcomes in the LBC1936 using a robust systematic approach that closely aligns with diagnosing dementia in practice. Our methods go far beyond those used by many research studies, which lack the detailed medical data required to ascertain dementia in such a robust clinical manner. Previously, the best method available for determining dementia in the LBC1936 dataset was self-reported dementia status. This was included in LBC1936 as a sensitivity analysis to ensure the results of cognitive tests were not skewed by those who self-reported dementia. Therefore, it was heavily biased if used as a medical outcome for further research. We have illustrated in our comparison of self-reported and ascertained dementia outcomes that very few participants with dementia were well enough to communicate this information. Those more likely to drop out were at higher risk of dementia and ill-health.[3] The addition of this new dementia outcome using medical data linkage adds great value to the LBC1936 dataset.

In total, 118/865 (13.6%) participants met the criteria for a diagnosis of probable dementia between the ages of 69 and 85.5 years. The prevalence of dementia increased with age, and women had higher rates in nearly all age groups. The most common subtype was dementia due to Alzheimer disease (49.2%), followed by mixed Alzheimer and cerebrovascular disease (17.0%), dementia of unknown or unspecified cause (16.1%), and dementia due to vascular disease (8.5%).

### Comparison to literature

Our study’s all-cause dementia prevalence rates are comparable with other similar studies (community-based, neighbouring countries). For example, the prevalence rate for 75-79.9-year-olds in LBC1936 is 4.5% compared to two English cohorts Framingham (3.6%)[12] and the Cognitive Function and Ageing Studies (CFAS) II (5.2% [males] and 6.2% [females])[2], the male-only Caerphilly Prospective Study (3.9%)[13] from Wales, a cohort from Sweden (5.7%)[14], and a meta-analysis (5.6%)[15]. Additional File 3 presents the age-specific all-cause dementia prevalence rates across similar community cohorts from neighbouring countries.

Our finding of increasing prevalence with age and higher prevalence in women is common in most dementia prevalence studies.[2, 16–18] While the dementia rates in LBC1936 in the younger age groups (65-74.9 years) are low, the absolute numbers of dementia outcomes are small, so these prevalence rates should be interpreted cautiously. Similarly, we advise caution when interpreting the prevalence rate of our 85-89.9 years age group. At the time of our study, the mean age of participants with dementia in the 85-89.9 years group is only 85.49 years old. Previous meta-analyses found that dementia rates double every five years,[15, 16, 19] so it is reasonable to expect a large increase in dementia prevalence as the participants in the 85-89.9 years age group move towards the older end of the group over the next four to five years.

### Exploring variation

There are many difficulties with comparing prevalence rates in different studies using different methodological approaches. The diagnostic criteria for dementia and dementia subtypes have evolved since dementia population cohorts proliferated in the 1980s, making it especially difficult to compare estimates before this with newer ones.[20] Other methodological differences between studies also influence prevalence estimates. For example, in the Framingham study, only those scoring below set cut-off scores on the MMSE were called back for further evaluation, thus increasing the likelihood that they will have dementia.[12]

The slightly lower prevalence rates in our study may be partly explained by a trend towards reduced rates in later-born cohorts. In two landmark studies in cognitive ageing from England, CFAS I and II, there was a marked reduction in dementia prevalence rates over the 20 years between data collection instances.[2] Later-born populations had a lower risk of prevalent dementia than those born earlier in the 20^th^ century. This finding was replicated in a representative panel study, the English Longitudinal Study of Ageing (ELSA), which found a decrease in age-specific prevalence.[17] Despite this, the ELSA study reported that the number of people with dementia in England and Wales is projected to increase by 57% from 2016 to 2040, mainly due to improved life expectancy.[17]

Geography and socioeconomic status may also partly explain our slightly lower prevalence rates. The LBC1936 is a relatively healthy self-selecting cohort from a more affluent area than most in Scotland; their early life cognitive ability was higher, on average, than the general population,[4] and it may be that dementia rates are lower in Lothian than in other areas of the country.[3, 4] The CFAS I and II studies detailed important analyses of the effect of geography on dementia incidence and prevalence and found that prevalence varies according to deprivation indices in English localities.[2]

### Using mixed data sources

The proportion of Alzheimer disease among dementia outcomes in our study (49.2%) is comparable to the Framingham (55.6%)[12] and Kungsholmen (53.7%)[14] studies. Brain imaging results were available for 84% of the participants diagnosed with dementia. This was particularly important to subtyping vascular dementia, when brain imaging is especially helpful[9]. Dementia was noted on the death certificates of 73.4% of those with dementia who had died. This is similar to a previous Scottish study that found dementia was noted on the death certificates of 71.5% of patients who died with dementia.[21]

Using mixed data sources when ascertaining dementia outcomes is vital.[6] Our dementia ascertainment method of using a combination of existing data sources was validated in a study comparing diagnoses extracted from existing data with diagnoses made on clinical review in an earlier LBC cohort (LBC1921).[6] That study found that overall dementia diagnoses using data from multiple existing sources were confirmed by clinical review in 88% of outcomes.[6] A recent UK study found that using hospital admissions data alone unearthed 78% of dementia outcomes, and general practitioner data alone captured only 52% of dementia outcomes.[22] Ultimately, many dementias in the community remain undiagnosed as individuals affected do not attend health or social care services.[23] This makes it important that cohort studies have a system, like ours, of flagging individuals who merit clinical assessment for cognitive impairment, whether from concerns raised at the follow-up research waves, declining performance in cognitive tests, or some other warning sign.

### Strengths and Limitations

A major strength of this study is the limited attrition bias as we reviewed the EHR of all participants from wave 2 till the present (or their death). This is important, especially with an outcome like dementia, as participants with poorer cognitive ability are at a greater risk of loss to follow-up.[24] Several specialists were involved in reviewing the EHR and performing the home visits, and inter-rater variability was limited by having a clear protocol and a multidisciplinary consensus meeting including at least two people who had completed the EHR reviews. Our thorough EHR reviews combined with our system for flagging for home visit any participant presenting at wave 6 testing with evidence of cognitive dysfunction makes it very likely we captured anyone with concerns raised to the health service, or at LBC testing.

A limitation of the study was the inability to accurately provide subtypes for all those (n=118) who were diagnosed with dementia. This was mostly due to inadequate information recorded in the EHR for people who subsequently died (i.e., could not be assessed further by the study team). This reflects clinical practice in Scotland in the early 2000s, where subtypes were not always routinely recorded. Of note, no participants were diagnosed with frontotemporal dementia (FTD), whereas in recent dementia cohorts, FTD diagnoses have accounted for 1.6% to 6% of dementia outcomes.[25, 26] However, these cohorts tended to include relatively younger adults (mean age 64 years[26]). A further limitation is that the home visits were only for a small selection of our cohort based on a specific set of criteria (outlined in methods section) applied to those who attended the latest follow-up wave.

### Implications

The identification of whether an LBC1936 participant has dementia will be invaluable for future research identifying risk factors and associations with dementia in this well-characterised cohort. The LBC1936 has five waves (sixth is underway, seventh is planned) of consistently measured cognitive, brain imaging, biomedical, psychosocial, and lifestyle data covering the ages of 70 – 86 years. It has, uniquely, a measure of intelligence at age 11. The latest data types in LBC1936 include: whole-genome sequencing, longitudinal DNA methylation, longitudinal gene expression, lipidomics, post-mortem brain tissue, induced pluripotent stem cells, inflammatory markers, oxidative stress markers, life course geographical information, objectively measured physical activity and sedentary behaviour.[3] This ensures a vast range of possibilities for future dementia research.

## Conclusion

These dementia outcomes for the well-characterised LBC1936 can be a foundation for future studies to confirm existing, and assess novel risk factors for dementia, and contribute to the rational basis for the development of new interventions to reduce incident dementia.

## Supporting information

S File 1: EHR Protocol

S File 2: Person-hours

S File 3: Comparison table

## Data Availability

All data are available on reasonable request here: https://www.ed.ac.uk/lothian-birth-cohorts/data-access-collaboration

## Acknowledgements

The authors are very grateful to the LBC1936 participants past and present, and the LBC research team who recruited participants, obtained data linkage permissions and collected data. Their commitment and contributions have made this work and many other important studies possible. Dr Mullin is very grateful for the support of his funders, supervisors, and academic mentors. For the purpose of open access, the authors have applied a CC-BY public copyright licence to any Author Accepted Manuscript version arising from this submission.

## List of abbreviations

CFAS: Cognitive Function and Ageing Studies
CI: Confidence Interval
CT: Computerised Tomography
DSM-5: Diagnostic and Statistical Manual of Mental Disorders - Fifth Edition
DaT: Dopamine Transporter
EHR: Electronic Health Records
ELSA: English Longitudinal Study of Ageing
FTD: Frontotemporal Dementia
ICD-11: International Classification of Diseases-11
LBC1936: Lothian Birth Cohort 1936
MRI: Magnetic Resonance Imaging
MMSE: Mini-Mental State Exam
NHS: National Health Service
SPECT: Single-Photon Emission Computerised Tomography
SD: Standard Deviation

