## Supplementary material for "Dementia diagnosis and prevalence in the Lothian Birth Cohort 1936 using medical data linkage": S File 1: EHR Protocol

**Additional File 1**

**Dementia ascertainment protocol- Lothian Birth Cohort 1936 Study**

1. AT will securely email the participant list from LBC
2. Divide participants who have given consent between AS, CG and GW
3. For each participant search CHI/ UHPI number on TRAK
4. Search Trak diagnosis list (via clinical history, then previous codes) for coded diagnoses of dementia or cognitive impairment
5. Scan home appointment screen for MATS (Memory Assessment & Treatment Service) appointment- attended/ not attended/ booked
6. If MATS appointment- search SCI store/ correspondence for letter
7. Document certainty of diagnosis (no dementia, possible dementia, probable dementia), subtype diagnosis (probable/ possible descriptor), code (if app), date of diagnosis, cognitive assessment score and scale used, capacity (yes/ no/ uncertain), cognitive enhancer medication (no/ yes + name) > Sheet 2 of Excel Spreadsheet on shared drive
8. Copy and paste text relating to capacity assessment and summary of diagnostic rationale > Word Document on the shared drive (one for each participant indexed by LBC number as the filename)
9. Scan home appointment screen for other relevant appointments/ admissions- Old Age Psychiatry, Medicine of the Elderly, Cognitive Disorders Clinic (under Neurology)
10. Complete #7 and #8 for each as appropriate (if admission go through ‘Clinical Notes’ for that admission and search for Psychological Medicine entries)
11. If DNA/ booked- search SCI store for GP referral
12. Scan waiting list for relevant appointments- MATS, POA, MOE etc
13. Look at KIS data for dementia/ cognitive impairment diagnosis
14. Look at ECS data for cognitive enhancer medication
15. Scan correspondence for dementia/ cognitive impairment and/or terminal illness diagnosis. CTRL + F “dementia”, “cogn”, “memory”.
16. For each patient on the spreadsheet: confirm that Trak has been searched, enter dementia diagnosis (probable, possible, no), capacity (yes, no, uncertain), terminal illness (yes/no), additional notes e.g. queries for Study Medics, and an indication if further info is available in Sheet 2 of the Excel Spreadsheet and Word Document on the shared drive.
17. If the patient is deceased, go through the protocol as above. In addition, look at pathology results for brain biopsy if available and document findings in Word document.

N.B. This is for participants who gave consent for access to their medical records even if they are not being invited to wave 6. The above protocol will be followed similarly for those invited for wave 6 when this is recommenced in 2021.
