## Supplementary material for "Dementia diagnosis and prevalence in the Lothian Birth Cohort 1936 using medical data linkage": S File 2: Person-hours

|  | **Number of researchers involved** | **Approximate number of hours (averaged across researchers)** | **Total** |
| --- | --- | --- | --- |
| **Planning stage** | 4 | 2 | 8 |
| **Phase 1** | 6 | 80 | 400 |
| **Phase 2** | 5 | 4 | 20 |
| **Phase 3** | 9 | 6 | 51 |
| **TOTAL** |  |  | **489** |
