## Supplementary material for "Dementia diagnosis and prevalence in the Lothian Birth Cohort 1936 using medical data linkage": S File 3: Comparison table

Additional File 2: Comparable prevalence rates for all-cause dementia

| **Age group, years** | **Year** | **Country** | **65-69.9** | **70-74.9** | **75-79.9** | **80-84.9** | **85-89.9** |
| --- | --- | --- | --- | --- | --- | --- | --- |
| **Our study LBC1936^*^** | **2022** | **Scotland** | **-** | **0.8%** | **4.5%** | **9.5%** | **9.9%^^^** |
| Framingham[1] | 1992 | England | 0.9% | 1.8% | 3.6% | 10.5% | 23.7% |
| Caerphilly[2] (males) | 2008 | Wales | 0.8% | 3.9% | 9.4% | 15.4% | - |
| CFAS I[3] (males) | 1991 | England | 1.7% | 2.2% | 5.7% | 14.6% | 19.8% |
| CFAS I[3] (females) | 1991 | England | 2.0% | 2.9% | 7.4% | 13.9% | 26.5% |
| CFAS II[3] (males) | 2011 | England | 1.2% | 3.0% | 5.2% | 10.6% | 12.8% |
| CFAS II[3] (females) | 2011 | England | 1.8% | 2.5% | 6.2% | 9.5% | 18.1% |
| Kungsholmen[4] | 1991 | Sweden | - | - | 5.7% | 9.6% | 20.4% |

Note: LBC1936, Lothian Birth Cohort 1936; CFAS I & II, Cognitive Function and Ageing Study I & II.

*We pooled the two groups 65 to 69.9 years and 70 to 74.9 years to preserve anonymity due to sample distribution.

^At the time of our study, the mean age of participants with dementia in the 85-89.9 years group is only 85.49 years old. This explains the relatively low prevalence rate in this group.

[1]	Bachman DL, Wolf PA, Linn R, et al. Prevalence of dementia and probable senile dementia of the Alzheimer type in the Framingham Study. *Neurology* 1992; 42: 115–119.

[2]	Fish M, Bayer AJ, Gallacher JEJ, et al. Prevalence and Pattern of Cognitive Impairment in a Community Cohort of Men in South Wales: Methodology and Findings from the Caerphilly Prospective Study. *Neuroepidemiology* 2008; 30: 25–33.

[3]	Matthews FE, Arthur A, Barnes LE, et al. A two-decade comparison of prevalence of dementia in individuals aged 65 years and older from three geographical areas of England: results of the Cognitive Function and Ageing Study I and II. *Lancet (London, England)* 2013; 382: 1405.

[4]	Fratiglioni L, Grut M, Forsell Y, et al. Prevalence of Alzheimer’s disease and other dementias in an elderly urban population. *Neurology* 1991; 41: 1886–1886.
